# A quantum-enhanced precision medicine application to support data-driven clinical decisions for the personalized treatment of advanced knee osteoarthritis: development and preliminary validation of precisionKNEE_QNN

**DOI:** 10.1101/2021.12.13.21267704

**Authors:** Nima Heidari, Stefano Olgiati, Davide Meloni, Federico Pirovano, Ali Noorani, Mark Slevin, Leonard Azamfirei

## Abstract

**Background:** The clinical problem of knee osteoarthritis is that, although some novel therapies are safe and effective, the response is variable, and defining the features that predict individual response remains a challenge. Variational quantum-classical and quantum machine learning (QML) algorithms based on parameterized quantum circuits (PQC) are promising experimental technologies which can improve the efficiency of precision medicine clinical decision support systems (CDSS) based on real-world data stored in large unstructured databases. In this paper we tested a quantum neural network (QNN) application to support precision data-driven clinical decisions to select personalized treatments for advanced knee osteoarthritis.

**Methods:** Following patients’ consent and Research Ethics Committee approval, we collected clinico-demographic data before and after the treatment from 170 patients eligible for knee arthroplasty (Kellgren-Lawrence grade ≥ 3, OKS ≤ 27, Age ≥ 64 and idiopathic aetiology of arthritis) treated over a 2 year period with a single injection of microfragmented fat. Gender classes were balanced (76 M, 94 F) to mitigate gender bias. A patient with an improvement ≥ 7 OKS has been considered a Responder. We trained our QNN Classifier on a randomly selected training subset of 113 patients to classify responders from non-responders (73 R, 40 NR) in pain and function at 1 year. Outliers were hidden from the training dataset but not from the validation set. We ran our QNN Classifier on a IBM quantum simulator to reduce the error due to noise.

**Results:** We tested our QNN Classifier on a randomly selected test subset of 57 patients (34 R, 23 NR) including outliers. The No Information Rate was equal to 0.59. Our application correctly classified 28 Responders out of 34 and 6 non-Responders out of 23 (Sensitivity = 0.82, Specificity = 0.26, F1 Statistic= 0.71). The Positive (LR+) and Negative (LR-) Likelihood Ratios were respectively 1.11 and 0.68. The Diagnostic Odds Ratio (DOR) was equal to 2.

**Conclusions:** Preliminary clinical and technical results of a QNN Classifier tested on a relatively small knee osteoarthritis dataset show that quantum machine learning applied to data-driven clinical decisions is a promising technology. Our results need further research validation with larger, real-world unstructured datasets, and clinical validation with an AI Clinical Trial to test model efficacy, safety, clinical significance and relevance at a public health level.

## BACKGROUND

Applied research has shown that quantum computing applied to machine learning is a novel experimental digital medicine technology [Cordier, Sawaya, Guerreschi, and McWeeney, 2021] which can reduce computational complexity [Sengupta and Srivastava, 2021, Singh and Aarthi, 2021] and improve data-driven clinical decisions [Solenov, Brieler, and Scherrer, 2018].

A number of biologic therapies, including the injection of Microfragmented Adipose Tissue, have been shown to be safe and effective [Heidari, Noorani, Slevin, Cullen, Stark, Olgiati, Zerbi, and Wilson, 2020, Borg, Heidari, Noorani, Slevin, Cullen, Olgiati, Zerbi, Danovi, and Wilson, 2021, Heidari, Borg, Olgiati, Slevin, Danovi, Fish, Wilson, and Noorani, 2021a]. The outcomes of these treatments can be variable [Sihvonen, Paavola, Malmivaara, and Järvinen, 2013] with some patients showing dramatic improvements in their symptoms whilst others fail to respond [Borg, Heidari, Noorani, Slevin, Cullen, Olgiati, Zerbi, Danovi, and Wilson, 2021].

Defining the characteristics of individual who will respond remains a challenge. Applications and tools to support clinical decisions with predictive, data-driven, precision medicine can identify patients who will respond with reduction in pain and improvement in function to novel as well as established treatments based on pre-treatment clinico-demographic data [Heidari, Parkin, Olgiati, Meloni, Fish, Noorani, Slevin, and Azamfirei, 2021b]. This approach is key in developing personalized, evidence-based clinical pathways.

## METHODS

### Ethics Statement

This study was carried out in compliance with the rules of the Helsinki Declaration and International Ethical Regulations [Association et al., 2009], including all subsequent amendments, under the approval of the Research Ethics Committee of the George Emil Palade University of Medicine, Pharmacy, Science and Technology of Targu Mures, Romania (research approval number 1464/2021).

### Dataset

Following patients’ consent and Research Ethics Committee approval, we collected clinico-demographic data before and after the treatment from 170 patients eligible for knee arthroplasty (Kellgren-Lawrence grade ≥ 3, OKS ≤ 27, Age ≥ 64 and idiopathic aetiology of arthritis) treated over a 2 year period with a single injection of microfragmented fat. (Table 1)

**Table 1.** Baseline distribution of Age on Procedure, Pre-operative Oxford Knee Score, and Kellgren-Lawrence grade of knee osteoarthritis. Source Dataset 1464/2021

|  | Age on Procedure | Pre-operative OKS | KL Grade |
| --- | --- | --- | --- |
| mean | 74 | 21 | 4 |
| std dev | 7 | 6 | 0.4 |
| min | 65 | 0 | 3 |
| 25% | 69 | 18 | 4 |
| 50% | 72 | 21 | 4 |
| 75% | 77 | 25 | 4 |
| max | 92 | 27 | 4 |

Gender classes were balanced (76 M, 94 F) to mitigate gender bias.A patient with an improvement ≥ 7 OKS has been considered a Responder.

### Oxford Knee Score (OKS)

OKS comprises of 12 questions that are scored 0-4 with 0 being severe compromise and 4 being no compromise, covering pain and function of the knee. The best outcome is a score of 48 and the worst score possible is 0. This is a validated score or the measure of functional outcomes in patients undergoing knee arthroplasty. All 170 patients completed questionnaires before and at three months, six months and one-year following treatment [Dawson, Fitzpatrick, Murray, and Carr, 1998].

### Kellgren-Lawrence method of classifying the severity of osteoarthritis (OA)

The Kellgren and Lawrence system is a common method of classifying the severity of osteoarthritis (OA) using five grades [Kohn, Sassoon, and Fernando, 2016].

- grade 0 (none): definite absence of x-ray changes of osteoarthritis
- grade 1 (doubtful): doubtful joint space narrowing and possible osteophytic lipping
- grade 2 (minimal): definite osteophytes and possible joint space narrowing
- grade 3 (moderate): moderate multiple osteophytes, definite narrowing of joint space and some sclerosis and possible deformity of bone ends
- grade 4 (severe): large osteophytes, marked narrowing of joint space, severe sclerosis and definite deformity of bone ends

Osteoarthritis is deemed present at grade 2 although of minimal severity 1.

### Model and Circuit Description

We applied the approach to quantum problem solving developed by Zickert [2021]. We utilized a Parameterized Quantum Circuit (PQC) based on Quantum Machine Learning Neural Networks Classifier (QNN) technology developed by IBM and Qiskit Machine Learning. More specifically, we utilized OpflowQNN which is a neural network based application for the evaluation of quantum mechanical observables. OpflowQNN circuit is described in Fig. 1. where:

**Fig. 1.**
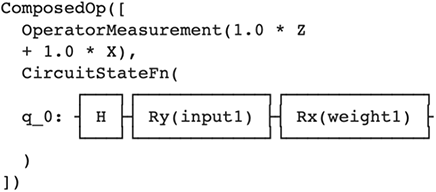
Circuit Description. Source: IBM, Qiskit and OpflowQNN.

- q_0 = qubit initialization
- Z, X = Pauli Z and X gates
- H = Hadamard gate
- Ry = single-qubit rotation gate through angle *θ* (radians) around the y-axis.
- Rx = single-qubit rotation gate through angle *θ* (radians) around the x-axis

### Quantum Simulator

The limit of existing quantum computers is quantum noise, i.e. the maximum computation time is measured in milliseconds rather than seconds (Noisy Intermediate-Scale Quantum, or NISQ) Bharti, Cervera-Lierta, Kyaw, Haug, Alperin-Lea, Anand, Degroote, Heimonen, Kottmann, Menke, et al. [2021]. This noise can induce further error in the estimate of the performance of our QNN Classifier Zickert [2021]. For this reason, we decided to utilize a IBM’s high-performance simulator locally. IBM [2021]

### Copyright Notice

Our code is an alteration of Qiskit Machine Learning which is a Copyright of IBM 2017, 2021. Qiskit is licensed under the Apache License, Version 2.0. All modifications of the code and derivative works are the responsibility of the authors.

### Model training

We trained our QNN Classifier on a randomly selected training subset of 113 patients treated over a 2 year period with a single injection of microfragmented fat classified as responders (R; OKS improvement ≥ 7) and non-responders (R; OKS improvement < 7) in pain and function at 1 year (73 R, 40 NR). Outliers were hidden from the training dataset but not from the validation set. (Fig. 2)

**Fig. 2.**
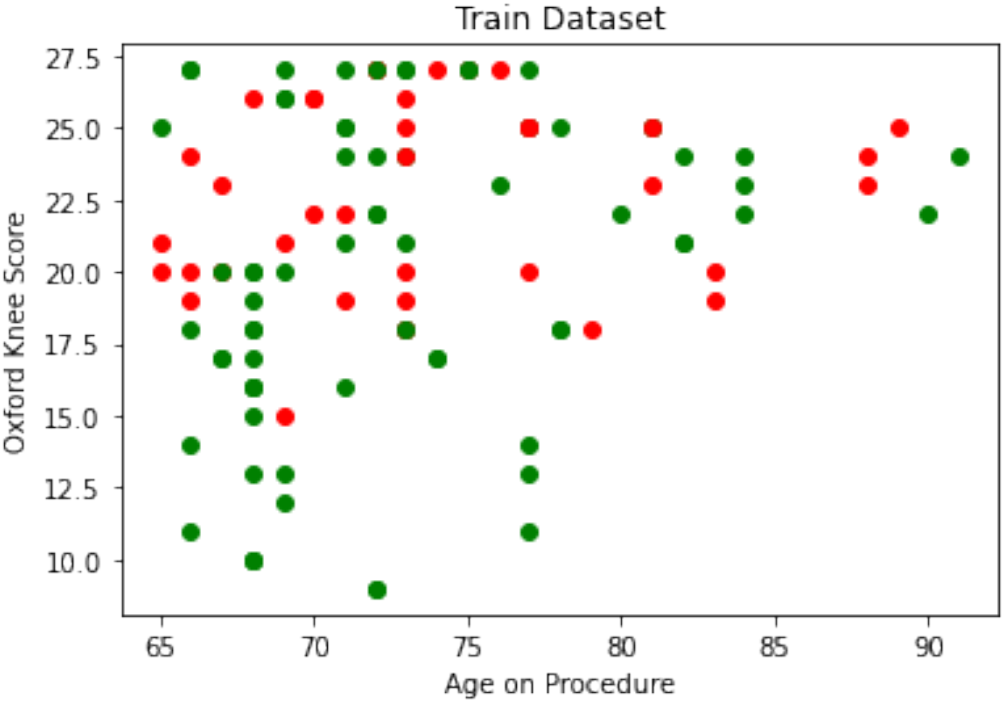
X-axis = Age on Procedure Y-Axis = Pre-operative Oxford Knee Score; Green dots = Responders with an improvement of ≥ 7 points in the Oxford Knee Score at Year 1; Red dots = Non-Responders with an improvement of < 7 points in the Oxford Knee Score at Year 1. Source Dataset 1464/2021.

We initialized a 2-qubit Aer Simulator Backend with 1024 shots. We utilized a “Constrained Optimization BY Linear Approximation” (COBYLA) optimizer to minimize the Objective Function Value. (Fig. 3)

**Fig. 3.**
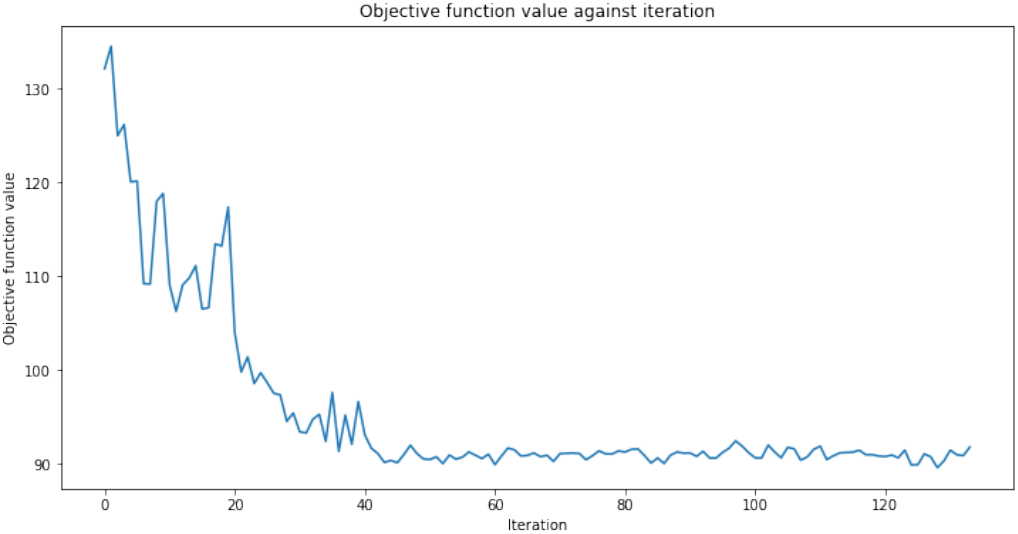
Constrained Optimization BY Linear Approximation11 (COBYLA) optimizer to minimize the Objective Function Value. Source: IBM, Qiskit and OpflowQNN.

### Model validation

The application has been tested on a balanced raw dataset including outliers of 57 patients (34 R, 23 NR) with Kellgren-Lawrence grade = 3 and 4, OKS ≤ 27, Age ≥ 64 and idiopathic aetiology of arthritis. (Fig. 4, Table 3)

**Fig. 4.**
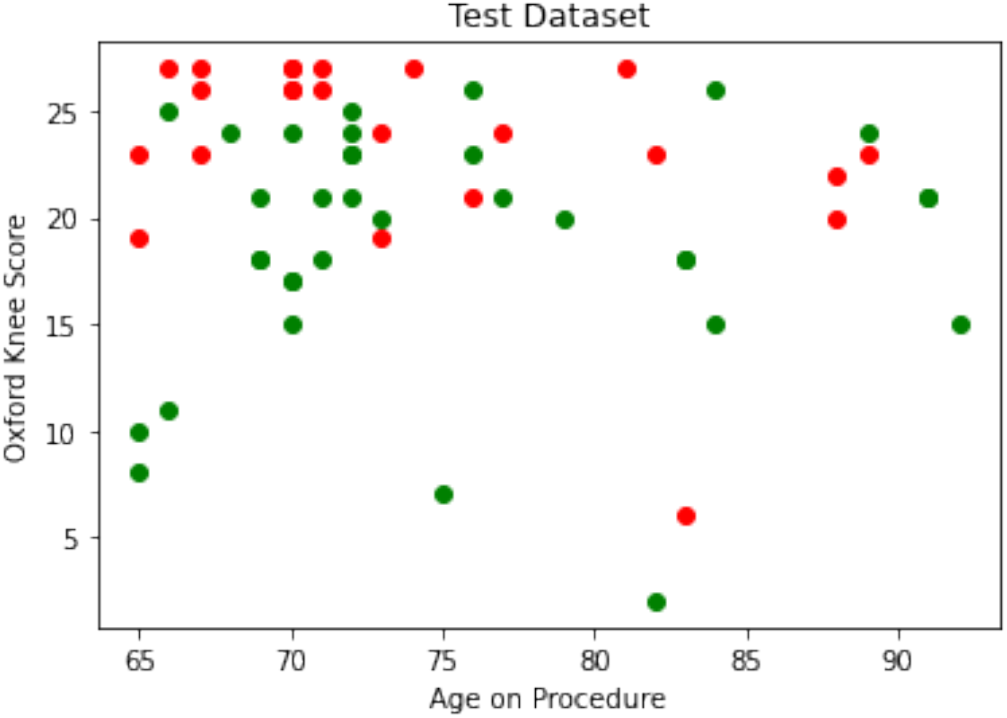
X-axis = Age on Procedure Y-Axis = Pre-operative Oxford Knee Score; Green dots = Responders with an improvement of ≥ 7 points in the Oxford Knee Score at Year 1; Red dots = Non-Responders with an improvement of < 7 points in the Oxford Knee Score at Year 1. Source Dataset 1464/2021.

## RESULTS

### Model Results

Our application correctly classified 28 Responders out of 34 and 6 non-Responders out of 23 (Sensitivity = 0.82, Specificity = 0.26, F1 Statistic= 0.71). The Positive (LR+) and Negative (LR-) Likelihood Ratios were respectively 1.11 and 0.68. The Diagnostic Odds Ratio was equal to 2. (Table 2 and Fig. 5)

**Table 2.**
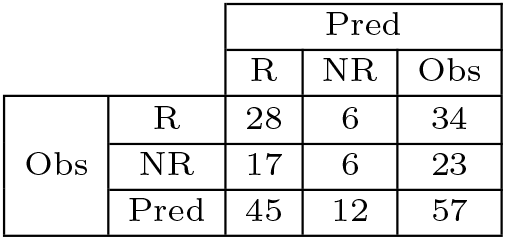
Confusion Matrix. Source precisionKNEE_QNN applied to Test Set from Dataset 1464/2021

**Table 3.** Test Performance Metrics. Source precisionKNEE_QNN applied to Test Set from Dataset 1464/2021

| Metric | Value |
| --- | --- |
| Sensitivity | 0.8235 |
| Specificity | 0.2609 |
| LR+ | 1.1142 |
| LR- | 0.6765 |
| DOR | 2.0000 |
| F1 | 0.7088 |

**Fig. 5.**
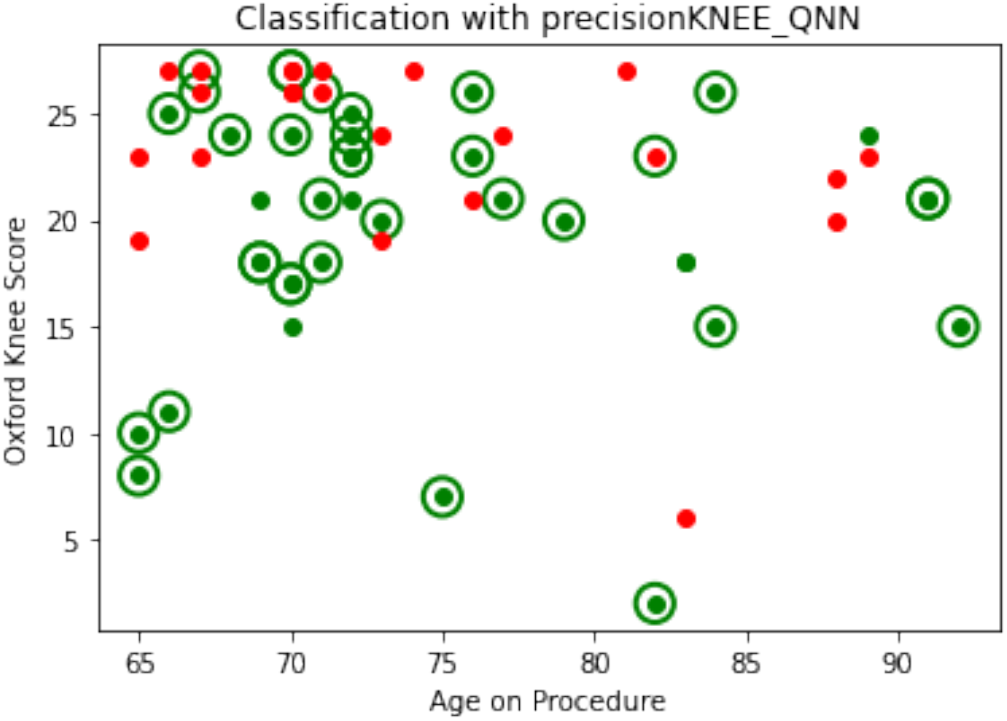
X-axis = Age on Procedure Y-Axis = Pre-operative Oxford Knee Score; Green dots = Responders with an improvement of ≥ 7 points in the Oxford Knee Score at Year 1; Red dots = Non-Responders with an improvement of < 7 points in the Oxford Knee Score at Year 1; Green Circles = Correct classification of either Responders or Non-Responders; No Circles = Incorrect classification of either Responders or Non-Responders. Source Dataset 1464/2021.

### Model Interpretation

The use of the precisionKNEE_QNN clinical decision-making tool requires external validation. This is a new technology best suited to large unstructured datasets. Some of traditional methods of validation may not be appropriate for quantum machine learning tools such as ours [Hidary, 2019].

The SPIRIT-AI [Rivera, Liu, Chan, Denniston, and Calvert, 2020]) and CONSORT-AI [Liu, Rivera, Moher, Calvert, and Denniston, 2020] initiatives provide guidance to improve the transparency and completeness of reporting of clinical trials evaluating interventions involving artificial intelligence. The utility of these initiatives is not clear for use in Quantum Machine Learning.

### Limitations

Despite being one of the largest datasets of its kind, upon stratification we observe groups of patients with minimal observations and thus we cannot conclude external validity in the findings pertaining to these groups. Kellgren-Lawrence classification of arthritis was found to be an important feature during modelling. This measure is crude and correlates poorly with patient symptoms.

MFAT is the only treatment used in our model. With inclusion of other treatment modalities, a true clinical decision-making support tool can be developed. Patients find these very useful and by providing a personalised prediction of the possible outcome, they will be able to play a more meaningful role in the decision-making process.

In addition, in order to reduce the error induced by quantum noise, we utilized a IBM high-performance quantum simulator; running precisionKNEE QNN on a real quantum computer would add even more error in the clinical decision support system.

## CONCLUSIONS

Preliminary results on a small validation dataset show that quantum machine learning applied to data-driven clinical decisions for the personalized treatment of advanced knee osteoarthritis is a promising technology that can improve computational efficiency and prognostic performance.

Our clinical and technical results need further research validation with larger, real-world unstructured datasets, and clinical validation with an AI Clinical Trial to test model efficacy, safety, clinical significance and relevance at a public health level.

## Data Availability

All data produced in the present work are contained in the manuscript

## AUTHOR CONTRIBUTIONS STATEMENT

S.O. and N.H. were responsible for conceptualization; S.O. and N.H. were responsible for methodology; S.O. and D.M. were responsible for the software; S.O., F.P., and N.H. were responsible for the formal analysis; N.H. and A.N. were responsible for the investigation; N.H. was responsible for the resources; N.H. was responsible for data curation; N.H. and S.O. were responsible for writing?original draft preparation; N.H., S.O., D.M. and F.P. were responsible for writing?review and editing; N.H. and S.O. were responsible for visualization. All authors have read and agreed to the published version of the manuscript. S.O., and N.H. are responsible for the algorithms and processes on bias mitigation.

## ETHICS STATEMENT

This study was carried out in compliance with the rules of the Helsinki Declaration and International Ethical Regulations, including all subsequent amendments, under the approval of the Research Ethics Committee of the “George Emil Palade” University of Medicine, Pharmacy, Science and Technology of Targu Mures, Romania (research approval number 1464/2021).

## ACKNOWLEDGMENTS

The authors acknowledge Angela Cullen as the Data Manager and Analyst.

